# Prevalence of Cardiovascular-Kidney-Metabolic Stages in US Adolescents and Relationship to Social Determinants of Health

**DOI:** 10.1101/2024.11.25.24317946

**Authors:** Carissa Baker-Smith, Abigail M. Gauen, Lucia C. Petito, Sadiya S. Khan, Norrina Bai Allen

## Abstract

**Importance:** Given that many risk factors for atherosclerotic cardiovascular disease (ASCVD) begin in childhood, knowledge of the prevalence of cardio-kidney metabolic syndrome (CKM) in adolescents and its risk factors is critical to understanding the etiology of ASCVD risk burden.

**Objective:** To calculate the proportion of US adolescents with CKM stages 0, 1, and 2 and to assess the social factors and behaviors most strongly associated with advanced CKM stage.

**Design:** Cross-sectional analysis of 2017-2020 US National Health and Nutrition Examination Survey (NHANES) sample data.

**Setting:** United States

**Participants:** Adolescents

**Exposure:** Social determinants of health, including family income to poverty ratio, health insurance, routine healthcare access, and food security, as well as behaviors including smoking, physical activity, and diet.

**Main Outcomes and Measures:** The prevalence of CKM stages 0, 1, and 2 in adolescents was measured using survey-weighted data. Generalized linear models were used to quantify associations between social factors, behaviors, and CKM staging.

**Results:** Of the 1,774 surveyed adolescents ages 12-18 years, representing 30,327,145 US adolescents, 56% (95% CI 52-60%) had CKM stage 0, 37% (33-40%) had CKM stage 1, and 7% (5-9%) had CKM stage 2. Physical activity score (1 to 100, 100=highest) was lowest among adolescents with CKM stage 2 (physical activity score for CKM 0: 60 (31), CKM 1: 60 (32), and CKM 2 49 (33); p=0.025). Other health behaviors, such as the DASH diet and nicotine scores, did not differ according to the CKM stage (p=0.477 and p=0.932, respectively). According to sex, race, ethnicity, and age-adjusted multivariate logistic regression analyses, a ratio of income to poverty level >1.85, having health insurance, and food security, were associated with a 32% (OR 0.68 [95% CI:0.52,0.89]), 40% (OR 0.60 [95% CI: 0.37, 0.99]), and 45% (OR 0.55 [95% CI: 0.41,0.73]) lower odds of CKM stage 1-2, respectively. After adjustment for all sociodemographic factors, only food security was associated with 41% lower odds of CKM stage 1-2 (OR 0.59 [0.43, 0.81]).

**Conclusions and Relevance:** CKM stage 1-2 in adolescents is most strongly associated with food insecurity.

Improved access to healthy food and policies to address food security may help prevent higher CKM stage, beginning in adolescence.

**Clinical Perspective:** 

**Key Points:** <u>Question</u>: What is the prevalence of cardio-kidney metabolic syndrome (CKM) in adolescents, and what social determinants of health factors are associated with CKM stages 0,1, and 2 in adolescents?

<u>Findings</u>: In this population-based study, 44% of adolescents are in CKM stages 1 and 2, and the presence of CKM stages 1 and 2 in adolescents is most strongly associated with lower household food security.

<u>Meaning</u>: By addressing social factors, such as food security in the US, we may improve the cardio-kidney metabolic health of adolescents and improve cardio-kidney metabolic health across the lifespan.

## Introduction

The cardiovascular-kidney metabolic (CKM) syndrome was first proposed by the American Heart Association in 2023 to describe the intersection and multidimensional relationship between cardiovascular, kidney, and metabolic disorders that lead to increased atherosclerotic disease (ASCVD) risk.^1,2^ Traditionally, obesity, hypertension, dyslipidemia, diabetes, lifestyle factors (diet, exercise), sleep, and tobacco use have been considered independent risk factors for ASCVD.^3^ However, health factors intersect and contribute to ASCVD. According to the CKM framework, the intersection of obesity, hypertension, dyslipidemia, and diabetes presents an even greater risk for ASCVD.^1,2^

CKM syndrome reflects a disease process that begins with dysfunctional adiposity (overweight or obese status), leading to changes in vascular, renal, and metabolic health. CKM syndrome ultimately leads to an increased risk for premature coronary artery disease. CKM stages include Stage 0 (no CKM risk factors), Stage 1 (presence of excess or dysfunctional adiposity), Stage 2 (presence of metabolic risk factors, including hypertriglyceridemia, hypertension, diabetes), Stage 3 (subclinical ASCVD) and Stage 4 (clinical CVD). Regression in staging is possible with substantial lifestyle change and significant weight loss.

Individual lifestyle choice may be an important determinant of CKM stage. However, multiple levels of influence often impact lifestyle choices, including interpersonal, community, and societal factors. Further, beyond behavioral and biological, there are physical/built environment, socio-cultural, and healthcare system domains of influence that impact health outcomes.^4,5^ In short, these social determinants of health (SDOH) may have a more significant impact on the CKM stage in adolescents than individual choice alone. Factors such as healthcare access, healthy food sources, food security, and household income may influence lifestyle and choice.^5^ A recent cross-sectional study of adult data from 3101 US counties assessing the relationship between socioenvironmental determinants of health (e.g., primary health care access, food security, rurality) and age-adjusted mortality (aaMR) attributable to CKM found that the aaMR attributable to CKM was positively associated with food security and fine particulate matter air pollution (PM2.5) and negatively associated with household income or primary health care access rate.^6^ A previously published study described the relationship between SDOH measures and AHA Life’s Simple 7 cardiovascular health (LS7 CVH) score in adolescents, including the relationship between food security, income-to-poverty ratio, health insurance, availability of a routine place for health care, and the LS7 CVH score.^7^ This study identified a strong relationship between SDOH and LS7 CVH score.

In this study, we describe the prevalence of CKM stages 0, 1, and 2 in a nationally representative sample of US adolescents and determine whether measures of SDOH are associated with CKM stage. Given that CKM staging extends across the lifespan, it is critical to consider the relationship between SDOHs and CKM stage in adolescents.

## Methods

The 22-item STROBE (Strengthening the Reporting of Observational Studies in Epidemiology) checklist was used to ensure transparency in this research. All data used in this study are publicly available at https://www.cdc.gov/nchs/nhanes/index.htm.

### Study Population

This study used data from the National Health and Nutrition Examination Survey (NHANES): 2017- 2020. NHANES is a large program conducted by the National Center for Health Statistics that collects demographic, socioeconomic, and health-related survey data every 2 years, except during the 2017-2020 pre-COVID-19 pandemic period. NHANES uses multistage probability sampling to identify an appropriate mix and number of participants from within the US. It oversamples underrepresented minorities to ensure that final participant percentages are representative of the US by age, race, and ethnicity. In this study, we included the most recent pre-COVID NHANES data and used data collected between 2017 and 2020 for persons 12 to 18 (immediate pre-COVID period). Our final sample included 1,774 adolescents, corresponding to a weighted sample of 30,327,145 US adolescents.

### Study Variables

#### Defining CKM Health in Adolescents

The AHA CKM framework was applied to classify adolescents into CKM stages 0, 1, or 2.^2^. We adapted previously provided definitions to match data available during routine NHANES visits (**Table 1**).^8^ As data on subclinical CVD is unavailable in adolescents in NHANES, the staging was truncated at stage 2. This is reasonable given that markers of subclinical ASCVD (e.g., coronary artery calcium scores, high sensitivity CRP (hsCRP), brain natriuretic peptide (BNP)) are not routinely employed in preventive pediatric cardiovascular clinical care and echocardiogram data (subclinical heart failure) is not collected as part of NHANES. Further, the likelihood of stage 4 CKM, as defined by clinical ASCVD in the absence of syndromes such as familial hypercholesterolemia and Kawasaki disease, remains highly unlikely and rare during adolescence.

**Table 1:** Determination of Cardio-Kidney Metabolic (CKM) Staging.

| CKM health stages | Definition<sup>1</sup> | NHANES mapping |
| --- | --- | --- |
| Stage 0: No CKM health risk factors | Individuals without overweight/obesity, metabolic risk factors (hypertension, hypertriglyceridemia, MetS<sup>2</sup>, diabetes), CKD, or subclinical/clinical CVD | 1. BMI <85<sup>th</sup> percentile<br>2. SBP and DBP <95<sup>th</sup> percentile<br>3. TG <150 mg/dL<br>4. No MetS<sup>2</sup><br>5. No self-reported diabetes<br>6. BG <126 mg/dL<br>7. HbA1c <6.5%<br>8. No CKD<sup>3</sup><br>9. No CVD<sup>4</sup> |
| Stage 1: Excess and/or dysfunctional adiposity | Individuals with overweight/obesity, abdominal obesity, or dysfunctional adipose tissue, without the presence of other metabolic risk factors or CKD<br>BMI ≥25 kg/m<sup>2</sup> (or ≥23 kg/m<sup>2</sup> if Asian ancestry*) and/or<br>fasting blood glucose ≥100–124 mg/dL or HbA1c between 5.7% and 6.4% | 1. BMI ≥ 85<sup>th</sup> percentile, fasting blood glucose between 100 and 124 mg/dL, or HbA1c between 5.7% and 6.4%<br>2. TG <150 mg/dL<br>3. No MetS<sup>2</sup><br>4. No self-reported diabetes<br>5. BG <126 mg/dL<br>6. HbA1c <6.5%<br>7. No CKD<sup>3</sup><br>8. No CVD<sup>4</sup> |
| Stage 2: Metabolic risk factors and CKD | Individuals with metabolic risk factors (hypertriglyceridemia (≥135 mg/dL), hypertension, MetS<sup>2</sup>, diabetes) or CKD | 1. TG ≥135 mg/dL, SBP or DBP ≥95<sup>th</sup> percentile, MetS<sup>2</sup>, self-reported diabetes, BG ≥126 mg/dL, HbA1c ≥6.5%, or CKD<sup>3</sup><br>2. No CVD<sup>4</sup> |

BMI indicates body mass index; CKD, chronic kidney disease; CKM, cardiovascular-kidney-metabolic; ASCVD, atherosclerotic cardiovascular disease; MetS, metabolic syndrome; SBP, systolic blood pressure; DBP, diastolic blood pressure; GFR, glomerular filtration rate; TG, triglycerides; BG, fasting blood glucose. \*There may be individual differences in goal BMI, but a threshold of <23kg/m<sup>2</sup> was adopted by the AHA CKM Presidential Advisory committee.
NHANES adolescents were mapped to the stage in which they met each criterion.
<sup>1</sup>Definitions according to AHA's definitions of CKM syndrome stages.<sup>2</sup>
<sup>2</sup>MetS defined as the presence of 3 or more of the following: (1) HDL cholesterol <40 mg/dL for males and <50 mg/dL for females; (2) triglycerides ≥150 mg/dL; (3) elevated blood pressure (SBP or DBP ≥ 95<sup>th</sup> percentile); and (5) fasting blood glucose ≥100 mg/dL.
<sup>3</sup>CKD defined as GFR < 30. GFR was calculated from height and serum creatinine using Bedside Schwartz equation.<sup>28</sup>
<sup>4</sup>CVD defined as participants ever having been told they had coronary heart disease, heart failure, stroke.
BMI percentiles were calculated using height (cm), weight (kg), age (years), sex, and BMI via the 'get\_BMIpercentile' function within the 'PAutilities' package in R (https://cran.r-project.org/package=PAutilities). SBP and DBP percentiles were calculated using BP, age (months), sex, and height (cm) via the 'p\_bp' function within the 'pedbp' package in R.<sup>29</sup>

Body mass index (BMI) percentiles for age and sex were included in the BMI score. For the blood pressure scores, the average of 2 systolic blood pressures (SBPs) and 2 diastolic BPs (DBPs) per person were used to generate SBP and DBP percentiles for age and sex.^9^ Laboratory tests directly measured cholesterol and fasting serum glucose.^10^ A diet score was determined based on quantiles of dietary approaches to stop hypertension (DASH) diet adherence.^8^ Physical activity was scored based on self-reported minutes of moderate-vigorous recreational activity per week. Nicotine exposure was scored according to self-reported use of tobacco. Sleep was not included due to the lack of sleep information in adolescents within NHANES.

#### Sociodemographic Determinants of Health

SDOH factors evaluated in this analysis covered 4 domains: household poverty, source of health insurance, access to health care, and food security. Household poverty level (poverty versus non-poverty) was calculated as the ratio of monthly family income to poverty levels defined by US Department of Health and Human Services guidelines. A ratio of greater than 1.85 was used to create a binary variable based on standard eligibility guidelines for federal programs.^7^ Health insurance was categorized as having health insurance coverage, including private and public, vs. no coverage. Participants self-reported whether they had a routine place for health care available. The US Food Security Survey Module in NHANES includes 10 items for households without children and 18 for households with children. Data from households with children was analyzed. Households received a score of 1 if the household had complete food security (no affirmative responses to any of the 18 items), 2 if the household had marginal food security (1-2 affirmative responses), 3 if the household had low food security (3-7 affirmative responses), and 4 if the household had very low food security (8-18 affirmative responses). We defined food security as total or marginal food security, corresponding to a score of 2 or less.

We created a composite SDOH score by assigning 1 point for each favorable SDOH variable (ratio of income to poverty level >1.85, food security, health insurance, and having a routine place for health care).^7^ Each variable was weighted equally, with the SDOH score ranging from 0 (no favorable SDOH) to 4 (all favorable SDOH).

Last, demographic factors, including age (in years), sex (male, female), and race (non-Hispanic White, non-Hispanic Black, Hispanic, Other) were included as provided in NHANES. Race and ethnicity are according to self-report. Mexican Americans mainly represent the Hispanic population in NHANES. The “Other” adolescent classification included mixed-race adolescents and adolescents not included in the specific racial and ethnic categories.

### Statistical Analysis

The sociodemographic characteristics of the adolescents were described and stratified by CKM stage. Proportions were calculated for categorical variables, while means and standard deviations were calculated for continuous variables. Differences across the three CKM stages (e.g., 0, 1, 2) were estimated via Chi-Squared tests with Rao-Scott second-order corrections for proportions. Weighted univariable linear regression models with Wald tests for were used to assess differences across the three CKM stages for continuous variables.

Linear associations were estimated using weighted univariable linear regression models with continuous CKM stage. NHANES sample weights were used in all calculations; standard errors were estimated using Taylor series linearization.

Next, the associations between individual-level SDOH factors and CKM stages were estimated. A composite outcome variable of CKM stage 1-2 was constructed due to the relatively small sample size of adolescents with CKM stage 2. Sequential logistic regression models were performed to assess the associations between SDOH factors and CKM stage (1+2 versus 0) where M1 was the unadjusted model for each SDOH factor separately, M2 was M1 + age, sex, race, and ethnicity, and M3 included all SDOH factors. It was adjusted for age, sex, race, and ethnicity. Secondary analysis was performed where odds of higher CKM staging were estimated for the composite SDOH score, adjusted for age, sex, race, and ethnicity.

All analyses were performed using R version 4.3.0. Survey statistics were employed to provide population mean, proportion, and model estimates using the ‘survey’ package.^11^ Variables with missing data (<10%) were imputed via multiple imputations by chained equations using a package in R‘^12^; continuous and categorical data were imputed by predictive mean matching. Results were pooled over 20 imputed datasets using Rubin’s rules.^13^ Statistical significance was interpreted at p<0.05.

## Results

The sample included 1,774 adolescents, representing 30,327,145 adolescents, 12 to 18 years of age. Fifty-six percent (95% CI 52-60%) of adolescents had CKM stage 0, 37% (33-40%) had CKM stage 1, and 7% (5-9%) had CKM stage 2 (**Table 2**). The proportion of males with CKM stage 2 exceeded females (67% vs 33%, p=0.011) (**Table 3**). CKM staging also varied according to self-identified race and ethnicity (p=0.045). Of those with CKM stage 0, 52% were non-Hispanic White, 13% non-Hispanic Black, 21% Hispanic, and 14% of Other race/ethnicity. In contrast, of those with CKM stage 2, a lower proportion were non-Hispanic White (43%) and a higher proportion Hispanic (36%) (p=0.045).

**Table 2:**
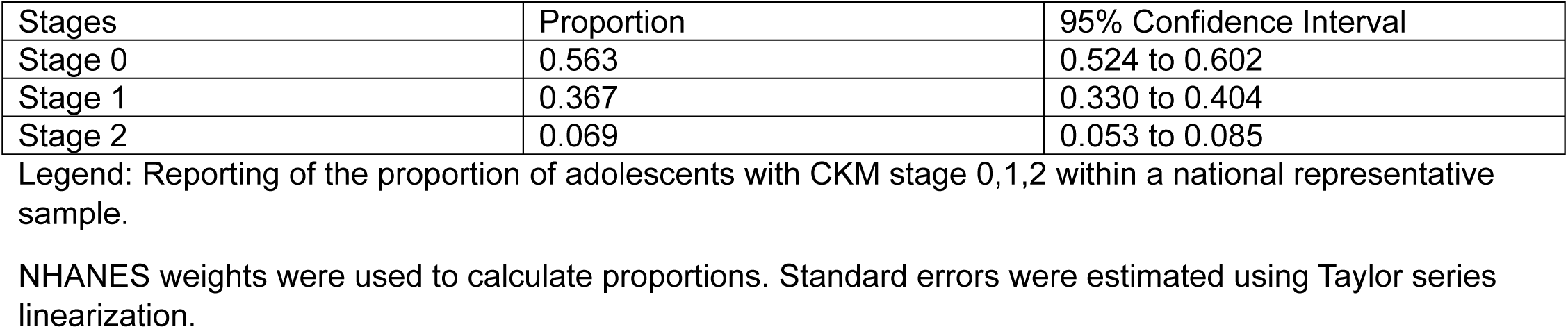
Proportion of Adolescents (Ages 12-18 years) in CKM Stages 0-2 in NHANES 2017-2020 (Weighted N=30,327,146)

| Stages | Proportion | 95% Confidence Interval |
| --- | --- | --- |
| Stage 0 | 0.563 | 0.524 to 0.602 |
| Stage 1 | 0.367 | 0.330 to 0.404 |
| Stage 2 | 0.069 | 0.053 to 0.085 |

Legend: Reporting of the proportion of adolescents with CKM stage 0,1,2 within a national representative sample.
NHANES weights were used to calculate proportions. Standard errors were estimated using Taylor series linearization.

**Table 3:** Demographics of the Pediatric Cardio-Kidney Metabolic (CKM) Patient by CKM Stage: NHANES 2017-2020 Adolescents, 12-18 years (Weighted N=30,327,145)

| Characteristic | Stage 0 | Stage 1 | Stage 2 | p-Value |
| --- | --- | --- | --- | --- |
| Weighted Total, N | 17380484 | 11551817 | 1394845 |  |
| Sex<sup>1</sup> |  |  |  |  |
| Female | 49% | 52% | 33% | 0.024 |
| Male | 51% | 48% | 67% |  |
| Age (years)<sup>2</sup> | 15.05 (2.03) | 14.77 (1.91) | 15.28 (1.92) | 0.055 |
| Race/Ethnicity<sup>1</sup> |  |  |  |  |
| Non-Hispanic White | 52% | 49% | 43% | 0.045 |
| Non-Hispanic Black | 13% | 15% | 11% |  |
| Hispanic | 21% | 27% | 36% |  |
| Other | 14% | 9% | 9.5 % |  |
| BMI (kg/m2)<sup>2</sup> | 20.1 (2.5) | 29.0 (5.7) | 30.8 (7.4) | <0.001 |
| Waist circumference (cm2)<sup>2</sup> | 72 (7) | 94 (13) | 100 (18) | <0.001 |
| Systolic blood pressure (SBP) (mm Hg)<sup>2</sup> | 108 (9) | 108 (9) | 115 (11) | 0.004 |
| Diastolic blood pressure (DBP) (mm Hg)<sup>2</sup> | 63 (7) | 65 (7) | 71 (8) | <0.001 |
| Hemoglobin A1c (%)<sup>2</sup> | 5.23 (0.24) | 5.26 (0.29) | 5.72 (1.22) | 0.002 |
| Blood glucose (mg/dL)<sup>2</sup> | 96.6 (4.2) | 97.2 (4.6) | 110.2 (34.2) | <0.001 |
| Triglycerides (mg/dL)<sup>2</sup> | 63 (21) | 75 (27) | 142 (57) | <0.001 |
| Total cholesterol (mg/dL)<sup>2</sup> | 152 (25) | 154 (28) | 171 (30) | 0.013 |
| High density lipoprotein (HDL-C) (mg/dL)<sup>2</sup> | 55 (11) | 48 (9) | 43 (11) | <0.001 |
| Glomerular filtration rate (GFR) (ml/min)<sup>2</sup> | 102 (21) | 103 (19) | 101 (22) | 0.212 |
| Diabetes<sup>1</sup> | 0% | 0% | 25% | <0.001 |
| Ratio of income to poverty level<sup>2</sup> | 2.75 (1.64) | 2.36 (1.58) | 2.05 (1.41) | 0.001 |
| Health insurance<sup>1</sup> | 95% | 91% | 88% | 0.047 |
| Routine place for health care<sup>1</sup> | 88% | 89% | 85% | 0.426 |
| Food security<sup>1</sup> | 86% | 76% | 72% | <0.001 |
| DASH diet score | 80 (8) | 71 (10) | 62 (10) | <0.001 |
| Physical activity score | 32 (27) | 34 (30) | 30 (28) | 0.477 |
| Nicotine exposure score | 60 (31) | 60 (32) | 49 (33) | 0.025 |

Legend: Categorical analyses via Chi-Squared test with Rao-Scott second-order correction. Continuous analysis via Wilcoxon rank-sum test for complex survey samples. Food security defined as $\leq 2$ affirmative responses to US Food Security Survey Module. American Heart Association (AHA) Life's Essential 8 (LE8).
<sup>1</sup>Weighted %
<sup>2</sup>Weighted mean (SD)
<sup>3</sup>LE8 score calculated without sleep

As expected, due to the definition of each CKM stage (**Table 1**), the severity of each CVD health factor (BMI, waist circumference, blood pressure, HgbA1c, blood glucose, triglyceride, and total cholesterol level) was greater according to the CKM stage (**Table 3**). While the mean (SD) BMI was 20.1 kg/m^2^ (2.5 kg/m^2^) among adolescents with CKM stage 0, it was 29.0 kg/m^2^ (5.7 kg/m^2^) among those with CKM stage 1 and 30.8 kg/m^2^ (7.4 kg/m^2^) among those with CKM stage 2 (*p<0.001*). Similarly, values for waist circumference (*p<0.001)*, SBP (*p=0.004*), DBP (*p<0.001*), hemoglobin A1c (HgbA1c) (*p<0.001*), blood glucose (*p<0.001)*, triglyceride level (*p<0.001)*, and total cholesterol (*p=0.013*) increased with increasing CKM stage. High-density lipoprotein cholesterol (HDL-C) (*p<0.001*) decreased with increasing CKM stage.

Our primary focus for this study was to assess the relationship between SDOH factors and CKM stage in adolescence. We found that SDOH factors differed by CKM stage. The ratio of household income to poverty level was greatest among adolescents with CKM stage 0 vs stage 2 (2.75 [1.64] vs 2.05 [1.41]) (*p*=0.001).

While access to a routine place for healthcare did not vary by CKM stage, access to health insurance was lower among adolescents with CKM stage 2 vs stage 0 (88% vs 95%). Importantly, food security differed by CKM stage (*p<0.001)*: adolescents who were more food secure were more likely to have CKM stage 0 (86%) vs stage 2 (72%). Adherence to the DASH diet was greatest among adolescents with CKM stage 0 (80%) vs stage 2 (62%) (p<0.001). Adherence to recommended physical activity was no different among adolescents with CKM stage 0 (32%) vs stage 2 (30%), (p=0.477). Nicotine use was lowest (higher score) among adolescents with CKM stage 0 (60%) vs stage 2 (49%), (p=0.025).

Finally, we assessed the associations between individual-level favorable SDOH factors and a composite outcome of CKM stage 1-2 (**Table 4**). In separate unadjusted univariate analyses (M1), the ratio of income to poverty level >1.85, food security, and health insurance was associated with lower odds of CKM stage 1-2 (OR 0.66 [0.49, 0.87], 0.51 [0.38, 0.68], and 0.57 [0.34, 0.93], respectively). These associations remained after adjusting for age, sex, race, and ethnicity (M2) (OR 0.68 [0.52, 0.89], 0.55 [0.41, 0.73], and 0.60 [0.37, 0.99], respectively). After further adjustment for all four SDOH factors (M3), only food security was associated with lower odds of CKM stage 1-2 (OR 0.59 [0.43, 0.81]). Having a routine place for health care was not associated with lower odds of CKM stage 1-2 in M1, M2, or M3 (OR 1.05 [0.80, 1.38], 1.06 [0.84, 1.35], and 1.27 [0.96, 1.67], respectively). The composite favorable SDOH score was associated with lower odds of CKM stage 1-2 in both unadjusted and adjusted analyses (OR 0.76 [0.66, 0.87] and 0.77 [0.68, 0.86], respectively, for a one-point increase in SDOH score).

**Table 4:**
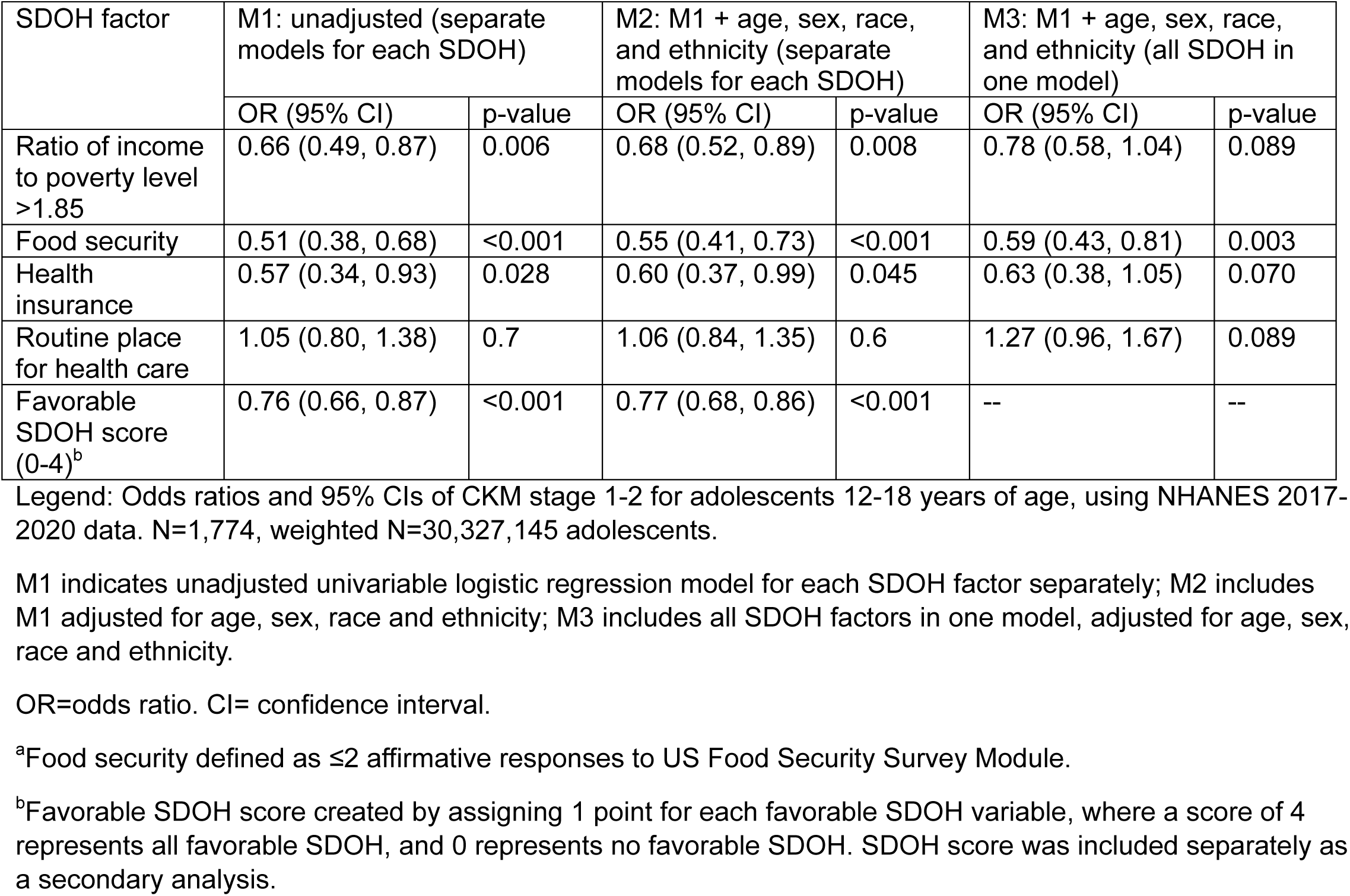
Sociodemographic Factors Associated with Cardio-Kidney Metabolic (CKM) Stage 1-2 vs Stage 0 in NHANES 2017-2020 Adolescents, 12-18 years (Weighted N=30,327,145)

## Discussion

Understanding the relationship between CKM stage and SDOH indicators in adolescents is important for managing CVD risk and reducing CKM and ASCVD risk across the lifespan. We found that roughly 44% of adolescents, 12 to 18 years of age, have CKM stage 1 or stage 2 and that the factor most strongly associated with CKM stage 1 or 2 was food security.

Consistent with prior published studies, our findings suggest that the prevalence of higher CKM stage increases with age. Aggarwal *et al.* found that roughly 75% of US adults 20 years of age and above, surveyed between 2011 and 2020 as part of NHANES, have Stage 1 or Stage 2 CKM.^14^ This is in contrast to our findings of 44% of adolescents with Stage 1 or 2 CKM. In this same study by Aggarwal *et al*, the prevalence of Stage 1- 2 CKM increased from 79.8% among adults 20-44 years to 83.9% among adults 45-61 years of age. By 65+ years of age the prevalence of Stage 1-2 CKM was surpassed by the prevalence of more advanced CKM stage (3-4) at 55%.^14^ Thus, the prevalence of more advanced CKM stage increases with age.

The presence of dysfunctional adiposity likely sets the stage for more advanced cardiometabolic disease. Despite the absence of clearly defined pathophysiologic mechanisms, composite risk factor development in childhood and adolescence is known to be associated with ASCVD in adulthood.^15^ Most adolescents with CKM stage above 0 have CKM stage 1 (37%): overweight/obesity status. Dysfunctional adipose tissue is metabolically active and leads to the development of diabetes, hypertension, and dyslipidemia. Inflammatory cells invade dysfunctional adipose, liver, and pancreatic tissue. Higher liver fat accumulation frequently develops, resulting in the development of metabolically active steatotic liver disease.^16^ Damage to the pancreatic beta cell leads to reduced insulin production and insulin deficiency. Further declines in beta cell dysfunction ultimately lead to the development of type 2 diabetes.^17^ Obesity-related elevations in serum leptin levels are associated with the development of hypertension.^18^ Ultimately, the composite of these risk factors leads to the development of ASVCVD, although the precise mechanisms by which inflammation contributes to greater ASCVD risk have not been clearly defined.^19^

Given that ASCVD risk is also influenced by SDOH, we assessed the relationship between SDOH and CKM stage during adolescence. The findings from our study are novel in that no study to date has assessed the relationship between food security and CKM stage in adolescents. We identified an important association: greater food security is associated with CKM stage 0 while lower food security (e.g., greater food insecurity) is associated with CKM stages 1 and 2 in adolescents. Like published findings in adults, we found that the more favorable the SDOH score, the more likely an adolescent was to have lower a CKM stage.^20^ However, our study went a step further. We identified the exact SDOH factors most associated with CKM stage 1 or 2 in adolescents. It was not the income-to-poverty ratio of the household, insurance coverage, or healthcare access that was mostly strongly associated with more CKM stage 1 or 2. Rather, food insecurity was the factor most associated with an adolescent developing CKM stage 1 or 2. The benefit of identifying this singular association after controlling for other SDOHs is that we can aptly identify appropriate interventions.

Food security in the United States has been a long-standing issue. The nation’s first attempts to address such deficits include the creation of the Food Stamp program in 1939 and the National School Lunch Program in 1946. However, approximately 25% of adolescents live in food-insecure households.^21^ Very low food security is prevalent in 5.1% of households.^22^ Approximately 1/3 of households below the Federal poverty line have food insecurity. In the US, food insecurity tends to be greatest among single parents as well as Black and Hispanic households.^22^ The NIH Nutrition Health Disparities Framework (NHDF) highlights the complex intersection of individual, interpersonal, community, and societal levels of influence that intersect with the biological, behavioral, built environment, socio-cultural, and healthcare system domains of influence contributing to nutrition-related health disparities.^23^ Interventions that target these multiple levels and domains of influence may be most successful in addressing the burden of food insecurity. Food insecurity also reflects inequitable housing and employment policies and patterns.^24^ Neighborhood characteristics, including a high density of fast food and limited access to affordable healthy foods, are known contributors to higher rates of childhood obesity.^25^ Further, race-based differences in the CKM stage may, in fact, be related to neighborhood characteristics, historic policies (e.g., redlining)^26^, societal norms, and limits of the built environment. Regardless of race or ethnicity, food security is the factor most strongly associated with advanced CKM in adolescents.

The relationship between higher CKM stages and SDOH has been described in adults. A systematic review assessing the characteristics and cardiovascular outcomes of the adult population with food insecurity residing within high-income countries revealed that 46%-58% are female, 11-68% have an education less than high school, and 37-58% are predominantly non-Hispanic White.^27^ According to this systematic review, adults with food insecurity have a 2.5 (95% CI 1.2-5.3) greater odds of obesity and a 2.2 greater odds of myocardial infarction.^27^

The CKM presidential advisory recommends that all persons, including adolescents, undergo SDOH screening, including food security screening.^2^ In our study, we have confirmed the importance of this recommendation. Greater efforts to resolve food insecurity across multiple levels and domains may lead to a healthier adolescent population.

Our analysis has limitations, including limited descriptors of race. “Other” race is a nebulous descriptor, and it is unclear if other subpopulations within the “Other” may be more susceptible to food insecurity. Next, we were limited to 4 out of the 6 previously described SDOH descriptors.^7^ The 2017-2020 NHANES does not include household reference marital status or education, as reported in earlier cohorts of NHANES. However, despite the absence of these 2 additional factors, we were able to characterize the significant relationship between SDOH factors, namely food security and CKM staging in adolescents 12 to 18 years of age.

In conclusion, applying the CKM construct to US population-based data demonstrates a high burden of CKM stage 1 or 2 in adolescents. Interventions such as improved food security may help to reduce the burden of CKM stage 1 or 2, beginning in adolescents.

Source of Funding: Dr. Baker-Smith is supported by Delaware INBRE P20GM103446-23S4.

## Data Availability

NHANES data is publicly available at https://wwwn.cdc.gov/nchs/nhanes/Default.aspx.

## Abbreviations

ASCVD: atherosclerotic cardiovascular disease
BMI: body mass index
CKD: chronic kidney disease
CKM: cardio-kidney metabolic
CVH: cardiovascular health
CT: computed tomography
DBP: diastolic blood pressure
GFR: glomerular filtration rate
HDL: high density lipoprotein
HF: heart failure
NHANES: national health and nutrition examination survey
OR: odds ratio
SBP: systolic blood pressure
SDOH: social determinants of health
TG: triglyceride

---

**Supplemental Table 1.** Quantification of Cardiovascular Health Using the Life’s Essential 8 score in Adolescents in NHANES 2017-2020.

| Domain | CVH Metric | Method of Measurement | Quantification of CVH Metric |  |
| --- | --- | --- | --- | --- |
| Health behaviors | Diet | Self-reported daily intake of a DASH-style eating pattern.<br>**Metric:** Quantiles of DASH-style diet adherence | **Points**<br>100<br>80<br>50<br>25<br>0 | **Quantile**<br>≥95th percentile (top/ideal diet)<br>75<sup>th</sup> – 94<sup>th</sup> percentile<br>50<sup>th</sup> – 74<sup>th</sup> percentile<br>25<sup>th</sup> – 49<sup>th</sup> percentile<br>≤24<sup>th</sup> percentile (bottom/least ideal) |
|  | Physical activity | Self-reported minutes of moderate or vigorous physical activity per week. For adolescents <18y, this was calculated by number of days in a week active at least 60 min, multiplied by 60 min. For individuals 18y, this was calculated by multiplying days/week vigorous recreational activities by minutes vigorous activities plus days/week moderate recreational activities multiplied by minutes moderate activity.<br>**Metric:** minutes of moderate or vigorous intensity activity per week | **Points**<br>100<br>90<br>80<br>60<br>40<br>20<br>0 | **Minutes**<br>≥420<br>360 – 419<br>300 – 359<br>240 – 299<br>120 – 239<br>1 – 119<br>0 |
|  | Nicotine exposure | Self-reported use of cigarettes or inhaled nicotine-delivery system<br>**Metric:** 100 cigarettes smoked in lifetime, smoked tobacco in past 5 days | **Points**<br>100<br>50<br>12.5 | **Status**<br><100 cigarettes in lifetime and hasn't smoked tobacco in past 5 days<br>≥100 cigarettes in lifetime but hasn't smoked tobacco in past 5 days<br>Smoked tobacco in past 5 days<br>Subtract 20 points for living with active indoor smoker in home (or 12.5 if score is 12.5) |

